# Motivationally salient cue processing measured using the Monetary Incentive Delay (MID) task with electroencephalography (EEG): A potential marker of apathy in Huntington’s Disease

**DOI:** 10.1101/2022.08.10.22278645

**Authors:** Marie-Claire Davis, Aron T. Hill, Paul B. Fitzgerald, Julie C. Stout, Kate E. Hoy

## Abstract

We explored the utility of the Monetary Incentive Delay (MID) task with concurrent encephalography (EEG) as a marker of apathy in people with Huntington’s disease (HD) as well as neurotypical controls. Specifically, we assessed between and within-group differences in the amplitude of the P300 and Contingent Negative Variation (CNV) event-related potentials as a function of motivational salience. In contrast to neurotypical controls, HD participants’ ERP amplitudes were not differentially modulated by motivationally salient cues (i.e., signalling potential ‘gain’ or ‘loss’) compared to ‘neutral’ cues. Difference waves isolating amplitude specific to the motivationally salient cues were calculated for the P300 and CNV. Only the difference waves for ERPs elicited by ‘gain’ cues differentiated the groups. The CNV difference wave was also significantly correlated with clinical measures of apathy and processing speed in the HD group. These findings provide initial support for the use of the MID with EEG as a marker of apathy in HD, and its potential as a sensitive outcome measure for novel treatment development.

## Introduction

Huntington’s disease (HD) is a life-limiting, progressive neurological disease caused by pathological expansion of the trinucleotide repeat (cytosine adenine guanine; CAG) in the *huntingtin (HTT*) gene on chromosome 4; the number of repeats correlating with the age of motor symptom onset and other clinical features (Pender and Koroshetz 2011, Podvin et al. 2019). Dysfunction and degeneration within the cortico-basal ganglia-thalamocortical (CBGT) networks contributes to the characteristic triad of motor, cognitive and psychiatric symptoms (Papoutsi et al. 2014, Ross et al. 2014, Ross et al. 2017). Apathy is a highly prevalent psychiatric symptom in HD (estimates range between 47-70%), which worsens with disease progression and is associated with major functional burden (Fritz et al. 2018, Jacobs, Hart, and Roos 2018, Martinez-Horta et al. 2016, Paoli et al. 2017). Similarly, of the cognitive impairments associated with HD, processing speed is the earliest cognitive deficit to emerge, and like apathy, worsens with disease progression (Paulsen et al. 2017). There are currently no evidence-based treatments for apathy or cognitive deficits, such as reduced processing speed, in HD (Anderson et al. 2018, Gelderblom et al. 2017).

There is increasing consensus that apathy occurring across a variety of neurological disorders reflects dysfunction within the CBGT networks and associated neurotransmitter systems, particularly dopaminergic pathways (Chong and Husain 2016, Lanctôt et al. 2017, Le Heron, Apps, and Husain 2017, Levy and Czernecki 2006, Levy and Dubois 2006, Roth et al. 2016). As such, the CBGT networks may be suitable biological targets for trialling clinical interventions for apathy in HD (Insel 2015). Demonstrating engagement of these biological targets, however, requires finding experimental paradigms that are both conceptually relevant to apathy and able to elicit measurable neurophysiological changes in these neuroanatomical regions of interest (Insel 2015).

Candidate experimental paradigms can be found by parsing the broader concept of apathy and, reciprocally, motivation, into neurocognitive subprocesses (Chong 2018, Husain and Roiser 2018, Kringelbach and Berridge 2016). At a neurocognitive level, research implicates abnormal effort discounting secondary to dysfunctional dopaminergic transmission in the aetiology of apathy (Chong 2018, Chong et al. 2017, Chong, Bonnelle, and Husain 2016, McGuigan et al. 2019, Atkins et al. 2020). Effort discounting refers to the devaluation of potential rewards as a function of the effort required to obtain them (Atkins et al. 2020, Chong 2018, Chong et al. 2017, Chong, Bonnelle, and Husain 2016, Chong and Husain 2016). How rewards and effort costs are perceived and subsequently valued, however, is influenced by the motivational salience of the rewards or outcomes on offer (Bromberg-Martin, Matsumoto, and Hikosaka 2010, Chong 2018, Olney et al. 2018). Motivationally salient cues signalling desired outcomes trigger dopaminergic transmission that adaptively directs cognitive resources and behaviour (Bromberg-Martin, Matsumoto, and Hikosaka 2010, Olney et al. 2018).

The Monetary Incentive Delay (MID) task is an extensively utilised functional magnetic resonance imaging (fMRI) paradigm that identifies and quantifies activity in neuroanatomical regions associated with the processing of motivationally salient cues (Knutson et al. 2000). Over 200 studies using the MID have confirmed that motivationally salient cues indicating a potential monetary ‘gain’ or ‘loss’, elicit greater neural activation than motivationally ‘neutral’ cues (i.e., cues with no potential gain or loss), in key regions of the CBGT networks implicated in apathy (i.e., striatum, insula, amygdala and thalamus) (Dugré et al. 2018, Knutson et al. 2000, Oldham et al. 2018, Wilson et al. 2018). Importantly, the fMRI version of the MID task has already been associated with abnormal ventral striatal activation in people with premanifest HD (Enzi et al. 2012) and Parkinson’s disease (du Plessis et al. 2018) and has correlated with increased apathy in people with schizophrenia (Kirschner et al. 2016, Simon et al. 2010). The MID is one of the tasks recommended by the National Advisory Mental Health Council Workgroup on Tasks and Measures for Research Domain Criteria (RDoC) for use in clinical research, and practice effects on the task are reportedly minimal (National Institute of 2016). Aside from sustained alertness, the cognitive and physical demands of the MID on participants is minimal, making it accessible to people with HD at various stages of their disease.

The MID has also been successfully adapted for use with electroencephalography (herein EEG) which is valuable because EEG provides much greater temporal precision and flexibility compared to MRI (Broyd et al. 2012, Donamayor, Schoenfeld, and Munte 2012, Pegg and Kujawa 2020, Pfabigan et al. 2014). Event-related potentials (ERPs) following cue presentation, specifically the P300 and Contingent Negative Variation, have been found to reliably differ according to the motivational salience of the cue (Hill et al. 2018, Novak and Foti 2015, Pfabigan et al. 2014, Zhang et al. 2017). The P300 (herein P3) is the third positive peak post-stimulus, emerges in frontoparietal midline electrodes, and varies according to a multitude of stable and acute factors (Luck 2014, Polich 2012). The broadest working hypothesis regarding the functional significance of the P3 suggests that it is associated with increased attentional focus to support memory storage via neural inhibitory processes (Polich 2012). The Contingent Negative Variation (CNV) is a spatially non-specific negative potential elicited when a warning stimulus signals the need to respond quickly to a target (Luck 2014). The timing of the CNV varies according to the delay between the cue and target (Luck 2014, Glazer et al. 2018). Functionally, the CNV is thought to be related to the modulation of motor preparation by motivation or effort, because it predicts the quality of the subsequent behavioural performance (e.g., faster response times) (Glazer et al. 2018). Given that the EEG version of the MID is both conceptually relevant to apathy and able to elicit measurable physiological changes in neuroanatomical regions of interest, we sought to investigate its utility in people with late premanifest and early manifest HD.

We hypothesised that the P3 and CNV ERPs in response to motivationally salient cues on the EEG version of the MID would be lower in people with HD relative to neurotypical controls. We also hypothesised that P3 and CNV amplitudes would significantly correlate with HD participants’ performances on clinical measures of apathy and processing speed.

## Materials and Method

### Participants

We recruited 20 neurotypical controls and 22 people with HD aged between 18 and 75 years as part of a randomised single session within-subjects study of non-invasive brain stimulation for apathy in HD conducted between February 2019 and August 2021 (Australian New Zealand Clinical Trials Registry (ANZCTR): 12619000870156). This study received ethics approval from the Alfred Health, Calvary Health Care Bethlehem, and Monash University Human Research Ethics Committees, and all participants gave their written informed consent.

This paper presents the results of the analysis of baseline data using the EEG version of the MID. The results of the non-invasive brain stimulation investigation will be presented in separate papers. All participants in the HD group were genetically confirmed to have the HD CAG expansion and were in the late premanifest or early manifest stages of their disease, defined as a disease burden score (DBS) of ≥280 (DBS = [CAG repeats − 35.5] × current age) and Total Functional Capacity score (TFC) of ≥7 (maximum score of 13) (Ghosh and Tabrizi 2018, Langbehn et al. 2004, Kieburtz 1996, Penney et al. 1997). The TFC is a clinician rating of how much assistance a person with HD requires to perform tasks in five functional domains that decline with disease progression (i.e., occupation, finances, domestic chores, activities of daily living, and care level) (Beglinger et al. 2010). A score of ≥7 corresponds with the early stages (i.e., stages I and II), during which the person with HD has motor symptoms, but can manage their domestic responsibilities and perform their usual activities of daily living (Ghosh and Tabrizi 2018, Shoulson and Fahn 1979). During stage II the person may have had to reduce their level of occupational engagement (e.g., work hours or responsibilities) and may require some assistance with managing their financial affairs (Shoulson and Fahn 1979, Ghosh and Tabrizi 2018).

Exclusion criteria included use of anticonvulsant medications or benzodiazepines; commencement or change in dose of other psychotropic medications (i.e., anti-depressants, anti-psychotics) during the four weeks prior to, and during participation; choreiform movements precluding EEG data collection; a current episode of psychiatric illness, or a current substance use or alcohol use disorder as assessed and defined by the Mini International Neuropsychiatric Interview (MINI) 7.0.2 (Sheehan et al. 1997); and a history of significant head injury or traumatic brain injury, as defined by a loss of consciousness greater than 30 minutes or requiring a hospital admission.

Eleven of the 22 participants with HD had been clinically diagnosed as having ‘manifest’ HD, the remaining CAG expanded participants were in the late premanifest stage (not yet diagnosed with motor symptoms but with a DBS ≥280) (Manifest group DBS M(SD) = 370.7(30.3), Range = 283.5-495.0; Late premanifest group DBS M(SD) = 351.0(60.3), Range = 286.0-484.5). Fourteen participants with HD were taking stable doses of psychotropic medications comprising SSRIs (n = 8), risperidone (n=7), mirtazapine (n = 3), SNRIs (n = 2), and tetrabenazine (n = 1). One of the control participants was on a low dose tricyclic antidepressant as a migraine prophylactic. Please refer to Table 1 for further demographic and clinical information.

**Table 1.** Characteristics of participant groups.

| **Table 1. Characteristics of participant groups** |  |  |  |  |
| --- | --- | --- | --- | --- |
| **Variable** | **Neurotypical controls**<br>**(n = 20)** | **HD CAG-expanded group**<br>**(n = 22)** | **Statistics** |  |
| Age in years (mean ±SD) | 47.40 (±13.90) | 48.59 (±11.23) | $t_{40} = -0.31$ | $p = 0.761^a$ |
| Range | 18-70 | 23-65 |  |  |
| Sex (male/female) | 9/11 | 10/12 | $\chi^2 = 0.001$ | $p = 0.976^b$ |
| Education years (median ±IQR) | 17.00 (±4.00) | 13.50 (±4.00) | $U_{42} = 77.50$ | $p < 0.001^c$ |
| Range | 13-21 | 11-20 |  |  |
| Work hours per week (median ±IQR) | 30.00 (±40.00) | 9.50 (±40.00) | $U_{42} = 179.50$ | $p = 0.289$ |
| Range | 0-50 | 0-50 |  |  |
| MoCA (median ±IQR) | 29.00 (±2.00) | 26.00 (±4.00) | $U_{39} = 45.50$ | $p < 0.001^c$ |
| Range | 25-30 | 19-29 |  |  |
| SDMT (mean ±SD) | 59.20 (±7.80) | 41.24 (±13.76) | $t_{39} = 5.10$ | $p < 0.001^a$ |
| Range | 44-74 | 14-71 |  |  |
| AES-C (median ±IQR) | 25.00 (±9.00) | 27.00 (±8.00) | $U_{41} = 306.50$ | $p = 0.011^c$ |
| Range | 13-35 | 23-53 |  |  |
| HADS – Anxiety (median ±IQR) | 4.00 (±2.00) | 5.00 (±5.00) | $U_{42} = 256.50$ | $p = 0.354^c$ |
| Range | 0-13 | 0-9 |  |  |
| HADS – Depression (median ±IQR) | 0.50 (±1.00) | 2.00 (±4.00) | $U_{42} = 339.50$ | $p = 0.002^c$ |
| Range | 0-3 | 0-10 |  |  |
| Psychotropic medication | 1 | 14 | - | - |
| CAG (median ±IQR) | N/A | 42.50 (±2.00) | - | - |
| Range |  | 40-52 |  |  |
| DBS in people with manifest HD (mean ±SD) | N/A | 370.7(30.3) | - | - |
| Range |  | 283.5-495.0 |  |  |
| DBS in people with late premanifest HD (mean ±SD) | N/A | 351.0(60.3) | - | - |
| Range |  | 286.0-484.5 |  |  |
| UHDRS-TMS (median ±IQR) | N/A | 11.00 (±11.00) | - | - |
| Range |  | 2-49 |  |  |
| Diagnosed as symptomatic by Neurologist | N/A | 11/22 | - | - |

<sup>a</sup>Independent samples t-test – 2-tailed; <sup>b</sup>Chi-Square test; <sup>c</sup>Wilcoxon rank sum/Mann Whitney U test; SD = standard deviation; IQR = inter-quartile range; MoCA = Montreal Cognitive Assessment (max score = 30); SDMT = written version of the Symbol Digit Modalities Test – written version (max score = 110); AES-C = Apathy Evaluation Scale – Clinician version (scores are between = 18 - 72); HADS = Hospital Anxiety and Depression Scale (subscale max scores = 21); CAG = number trinucleotide repeats on affected allele; DBS = Disease Burden Score (CAG-35.5 x age); UHDRS – TMS = United Huntington's Disease Rating Scale® - Total Motor Scale (max score = 131);

### Sample characterisation measures

The Unified Huntington’s Disease Rating Scale (UHDRS®) total motor score (clinician rated; Kieburtz 1996), Montreal Cognitive Assessment (MoCA) (rater administered; Nasreddine et al. 2005), Hospital Anxiety and Depression Scale (HADS) (self-reported; Zigmond and Snaith 1983), Apathy Evaluation Scale – clinician version (AES-C) (clinician rated; Marin, Biedrzycki, and Firinciogullari 1991), and Symbol Digit Modalities Test – written version (SMDT) (rater administered; Smith 1982) were administered as part of the study. The MoCA is a brief cognitive screen. The HADS screens for symptoms of anxiety and depression, with scores of ≤7 on both subscales considered to be within the ‘normal’ range (Snaith 2003, Stern 2014). The AES-C is a semi-structured clinical interview to assess the reductions in overt behaviour, emotional responsiveness and goal-directed thought content considered characteristic of apathy, with higher scores indicative of greater apathy (Marin, Biedrzycki, and Firinciogullari 1991). The SDMT is a timed, paper and pencil measure of processing speed that is sensitive, reliable, and well-validated for use in HD research (Stout et al. 2014). All sample characterisation measures were administered by MCD, an experienced clinical neuropsychologist with certification in the administration of the UHDRS® total motor score through the Enroll-HD clinical training portal (https://hdtraining.enroll-hd.org/). The UHDRS® total motor score and MoCA data were not collected for three HD participants, and SDMT and AES-C data are missing for one HD participant due to the COVID-19 social distancing restrictions and lockdowns in Melbourne Victoria, which impacted data collection for a small number of participants.

### Monetary Incentive Delay task

The version of the MID task we used was similar in timing parameters and stimuli to that used in previous research (Novak and Foti 2015, Pfabigan et al. 2014, Zhang et al. 2017) and was administered via Inquisit Lab version 4 software (Millisecond 2015). The task contained 10 baseline reaction time trials to establish the participant’s average response time; 12 practice trials (four of each gain, loss, and neutral cue type); and two blocks of 75 test trials (i.e., 150 test trials) comprising 50 gain cues, 50 loss cues, and 50 neutral cues, presented in random order. Gain cues signalled to the participant that they could win one dollar (+AU$1) if they pressed the enter key quickly enough after seeing a white square (target). If the participant was too slow, they did not win money. Loss cues signalled to the participant that they would *not* lose one dollar if they pressed the enter key quickly enough after seeing a white square. If the participant was too slow to respond to the white square, however, then they would lose one dollar (−AU$1). Neutral cues signalled to the participant that they were expected to respond as quickly as possible when they saw a white square but would neither gain nor lose any money during that trial. Feedback was provided after each response, and the participants’ cumulative winnings were updated at the base of the screen.

Each 7000 millisecond (ms) trial began with the cue presented for 1000ms, followed by an “anticipation phase” that randomly varied in 100ms increments between 2000-2500ms post-cue onset. The target (white square) was initially presented for a duration corresponding to the 60^th^ percentile of the participant’s average baseline response time. The target disappeared as soon as the participant responded. To ensure each participant obtained a comparable correct response rate of approximately 66% (Knutson et al. 2000), and to minimize floor and ceiling effects in participants’ response time data, the duration of target presentation increased by 30ms after 2 incorrect (i.e., too slow) responses, or decreased by 30ms after 3 correct (i.e., fast enough) responses using an adaptive algorithm (Hahn et al. 2009). Participants’ individualized target duration could increase to a maximum of 1500ms to accommodate slowed processing speed in participants with HD. Feedback was presented for 1000ms. The intertrial interval was at least 1000ms. Overall task duration was ~22 minutes. Participants were reimbursed a baseline sum of AU$10 but could earn up to an additional AU$30 per session, based on their winnings during the MID task, as per the standard administration guidelines to ensure task validity (Knutson et al. 2000). Refer to Figure 1 for stimuli and timing parameters.

**Figure 1.**
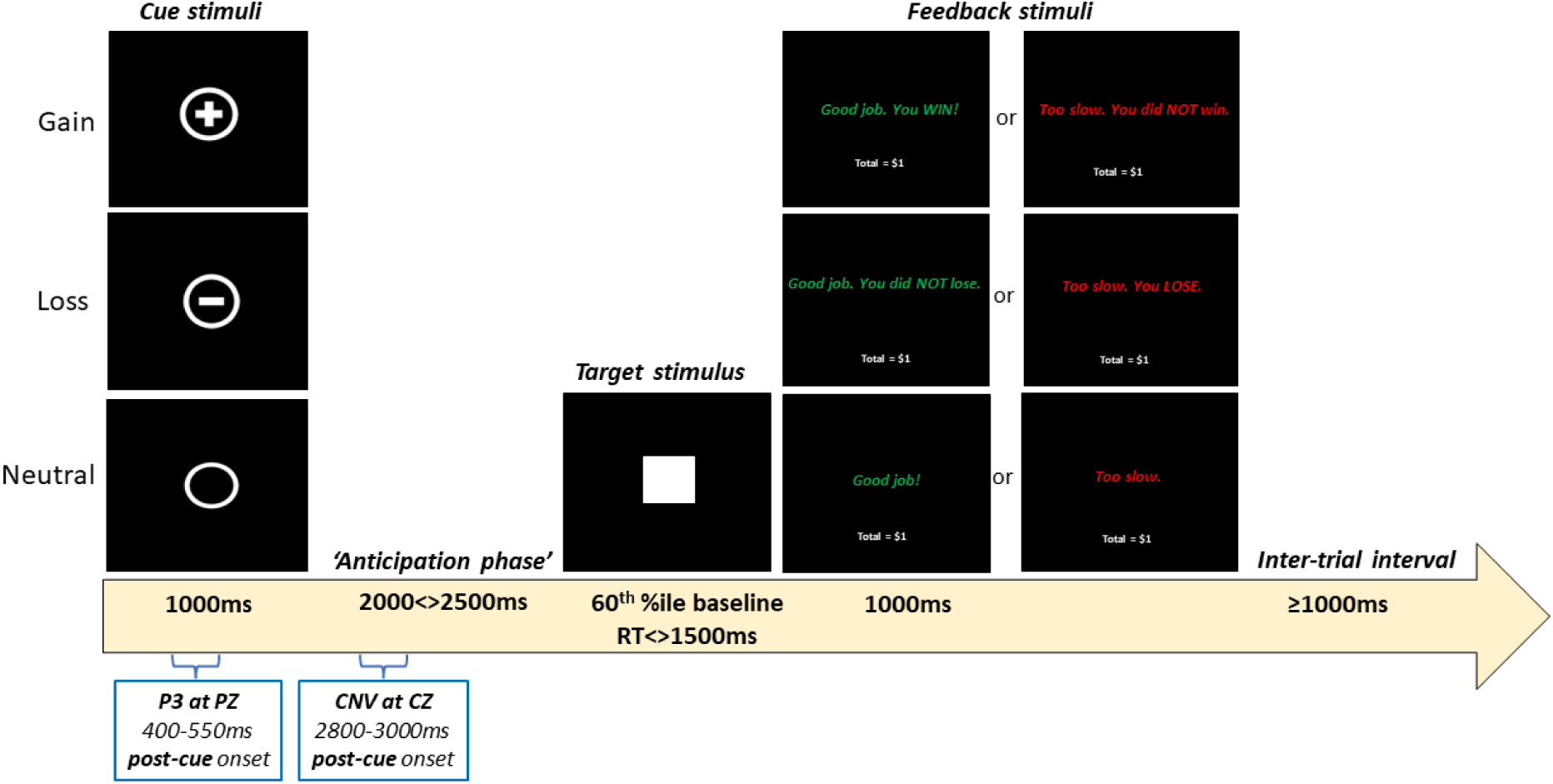
Stimuli and timing parameters for each 7000ms trial of the EEG version of the MID task

The P3 and CNV extracted from the MID data were based on the electrodes and timings from previous studies with similar stimuli and presentation timings (Broyd et al. 2012, Novak and Foti 2015, Pfabigan et al. 2014, Zhang et al. 2017). The P3 was comprised of the average amplitude from 400ms-550ms post-cue at electrode PZ, while the CNV was comprised of the average amplitude from 2800ms-3000ms post-cue at electrode CZ (refer to Figure 1). Difference waves were calculated, as described by Luck (2014), whereby the amplitude in response to neutral cues was subtracted from the amplitude in response to gain and loss cues.

### EEG recording and data processing

EEG was collected while participants completed the MID task using a 45-channel montage (EasyCap, Heersching, Germany) via the Neuroscan EEG system with Ag/AgCl electrodes connected to a SynAmps amplifier (Compumedics). Task presentation was synchronised with the EEG recording via the Inquisit 4 software, with EEG markers coinciding with the onset of each motivational cue presentation. The electrodes used were: AF3, AF4, F7, F5, F3, F1, FZ, F2, F4, F6, F8, FC5, FC3, FC1, FCZ, FC2, FC4, FC6, T7, C3, C1, CZ, C2, C4, T8, CP5, CP3, CP1, CPZ, CP2, CP4, CP6, P7, P5, P3, P1, PZ, P2, P4, P6, P8, PO3, POZ, PO4, O1, OZ, O2. Recording was completed at a sampling rate of 1KHz, with impedances kept below 5kΩ, and online referenced to CPZ with the ground at FCZ. The EEG data were pre-processed offline using RELAX, a fully automated EEG pre-processing pipeline (Bailey, Biabani, et al. 2022, Bailey, Hill, et al. 2022) through MATLAB (MathWorks 2019) and the EEGLAB toolbox (Delorme and Makeig 2004). Firstly, a notch filter (47<>52Hz) was applied to remove line noise, along with a fourth order Butterworth bandpass filter (0.25<>80Hz) with zero phase (Rogasch et al. 2017). The RELAX pipeline consisted of an initial extreme outlying channel and period rejection step, followed by a multi-channel Wiener filter algorithm which provided an initial reduction of eye movement, muscle activity and drift artifacts (Somers, Francart, and Bertrand 2018). Remaining artifacts identified via independent component analysis (ICA) using the automated ICLabel classifier were then removed using wavelet enhanced independent component analysis (wICA) (Pion-Tonachini, Kreutz-Delgado, and Makeig 2019). The initial bad electrode rejection was performed by the PREP pipeline (Bigdely-Shamlo et al. 2015), artifacts informing the Wiener filter algorithm were automatically identified using muscle artifact detection methods described by Fitzgibbon and colleagues (2016). The data were re-referenced to the average of all electrodes and segmented into non-overlapping 6.5-second epochs (−1.5 to 5.0 seconds around cue onset markers). Given that we were investigating ERP responses elicited by cues, all epochs were included for analysis, regardless of whether the participant’s response was ‘correct’ (i.e., they responded quickly enough on presentation of the white square). The number of epochs for each cue type remaining for analysis were as follows for neurotypical controls: Gain M(SD) = 42.13 (3.97); Loss M(SD) = 42.67 (3.73); Neutral M(SD) = 41.93 (3.83). The remaining epochs for participants with HD: Gain M(SD) = 42.08 (4.00); Loss M(SD) = 42.16 (4.00); and Neutral M(SD) = 41.86 (3.94). All epochs were baseline normalised from −500ms to −200ms prior to cue onset.

Using FieldTrip (Oostenveld et al. 2011), each participants’ epochs were averaged according to each cue type, with separate grand averages created for each cue type (gain, loss and neutral) for the HD and neurotypical control groups. The P3 and CNV ERPs for each cue type were extracted according to the previously specified parameters; namely, P3 was 400ms-550ms post-cue at electrode PZ, and CNV was 2800ms-3000ms post-cue at electrode CZ. Difference waves were calculated using FieldTrip (Oostenveld et al. 2011).

### Statistical analyses

Between-group differences in ERP amplitude and response time following each cue type (gain, loss and neutral) were assessed using mixed between-within subjects’ analysis of variance (ANOVA). Between-group differences in gain-neutral difference waves were analysed using t-tests. Participants’ average ERP amplitude values were extracted from those ERP analysis results that differed significantly between the HD and neurotypical control groups for exploratory correlation with the AES-C and SDMT. All statistical analyses were carried out using IBM SPSS version 28.0 (IBM Corp 2021).

## Results

### Group differences in clinical measures

As depicted in Table 1, the HD participants’ scores on the MoCA and SDMT were significantly lower than that of the neurotypical control group. The control group also had a significantly higher number of years of education than the HD group. Both apathy (AES-C) and the depression symptoms (HADS subscale) were significantly greater in the HD group than in the control group. Two participants with HD scored 8 and 10 respectively on the HADS depression subscale, putting them in the ‘mild’ (8-10) range for depression. Similarly, three neurotypical controls scored 8, 11 and 13 and one participant with HD scored 9 on the anxiety subscale, putting these participants in the ‘mild’ (8-10) and ‘moderate’ (11-14) ranges for anxiety on this screening measure (Stern 2014). Note however, that none of the participants from either group met criteria for a current psychiatric illness as assessed and defined by MINI.

### Behavioural responses

The groups did not differ significantly in their percentage of correct responses (*U_42_ = 245.000, p =0.523;* Median (IQR) HD = 61%(3%) and Control = 61%(3%)) indicating that the algorithm manipulating target presentation times was effective. Response times below 50ms were considered invalid and removed from further analyses (Pfabigan et al. 2014).

Between group differences in average response times to each cue type were then assessed using a Bonferroni adjusted mixed between-within subjects’ ANOVA. There was no significant interaction effect between participant group (HD vs control) and cue type (gain, loss, neutral) *(F_2, 39_ = 0.016, p = 0.98*). There were, however, significant main effects for both group (*F_1, 40_ = 13.37, p < 0.001, partial eta^2^ = 0.25)* and cue type (*F_2, 39_ = 4.24, p = 0.02, partial eta^2^ = 0.18)*. Compared to neurotypical controls, the average response times of people with HD were significantly longer following all three cue types. The differences in response time following each cue type were not significantly different (refer to Figure 2).

**Figure 2.**
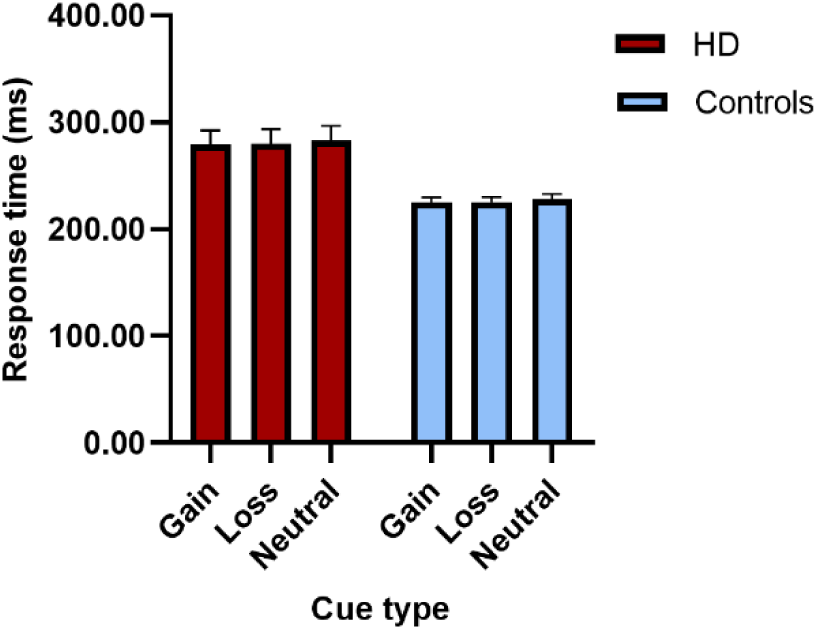
Response time in milliseconds (ms) for each group averaged across trials for the different cue types (error bars reflect standard error).

### P3 amplitudes

Between group differences in P3 amplitude at electrode PZ following each cue type were assessed using a Bonferroni adjusted mixed between-within subjects’ ANOVA. There was a significant interaction effect between participant group (HD vs controls) and cue type (gain, loss, neutral) (*F_2, 80_ = 4.48, p = 0.01, partial eta^2^ = 0.10*), as well as a significant main effect for cue type (*F_2, 39_ = 8.52, p < 0.001, partial eta^2^ = 0.30).* The main effect for group was not significant (*F_1, 40_ = 3.14, p = 0.08, partial eta^2^ = 0.07)*.

Post-hoc analyses were conducted to investigate the significant interaction effect. Differences between the groups based on cue type showed that the P3 amplitude for gain cues was significantly greater in the control group compared to the HD group (gain cue mean difference = 0.881, *p* = 0.02). The group differences in P3 amplitude for loss cues and neutral cues, however, were not significant (loss cue mean difference = 0.59, *p* = 0.10; neutral cue mean difference = 0.26, *p* = 0.42) (refer to Figure 3).

**Figure 3.**
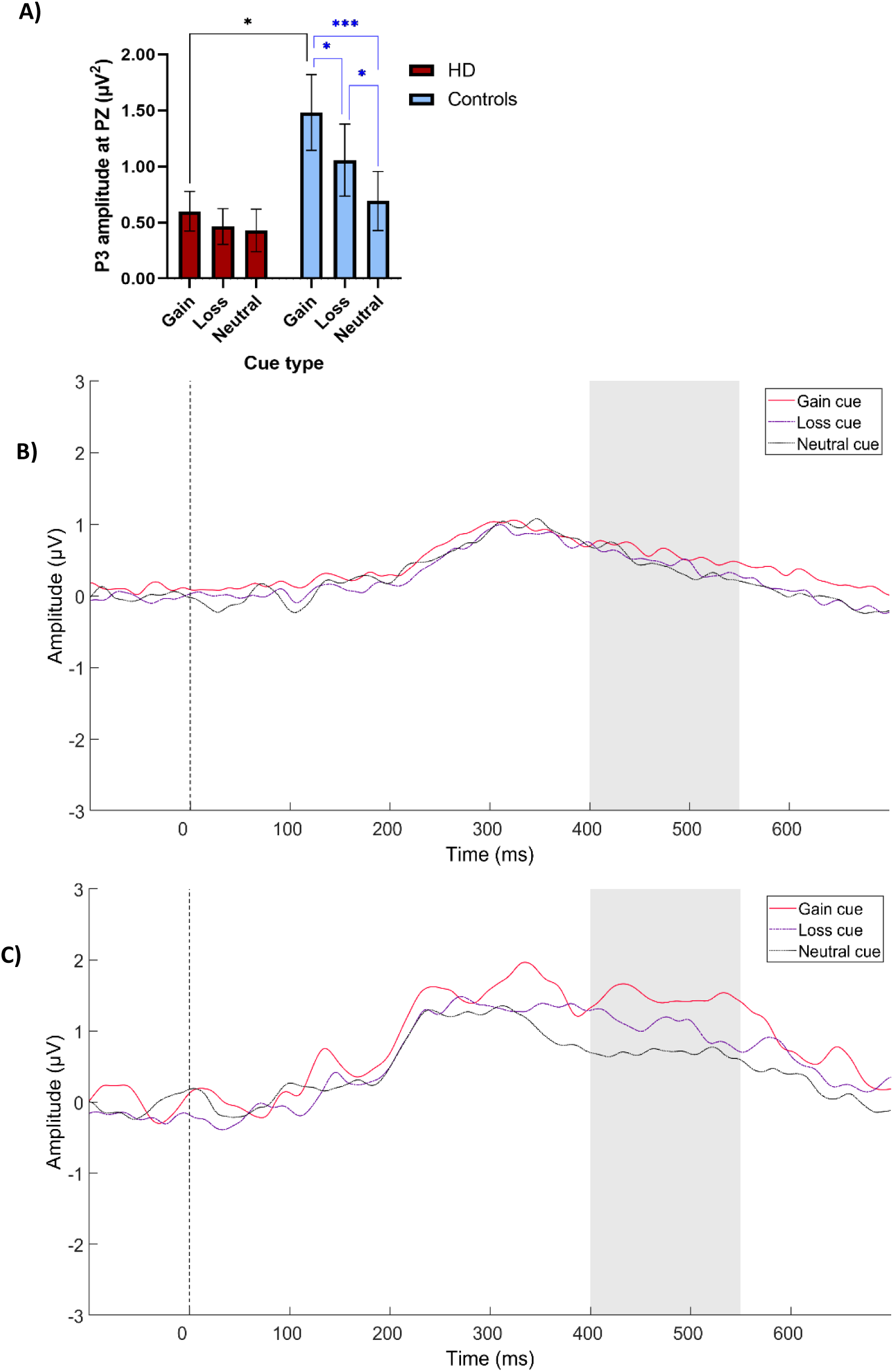
A) P3 amplitude at electrode PZ for each group averaged across trials for the different cue types (error bars reflect standard error). B) ERP plot of P3 at electrode PZ in response to each cue type for the HD group. C) ERP plot of P3 at electrode PZ in response to each cue type for the neurotypical control group. Grey bar indicates time window used for statistical analysis.

Pairwise comparisons of P3 amplitude in response to cue types within-groups revealed that in controls the P3 in response to gain cues was significantly greater than in response to loss cues (gain vs loss mean difference = 0.42, *p* = 0.02), which were in turn significantly greater than responses to neutral cues (loss vs neutral mean difference = 0.37, *p* = 0.01), with the largest difference between gain and neutral cues (gain vs neutral difference = 0.80, *p<0.001*). By contrast, for the HD group, there were no significant differences in P3 amplitude across the three cue types.

### CNV amplitudes

Between group differences in CNV amplitude at electrode CZ following each cue type were assessed using a Bonferroni adjusted mixed between-within subjects’ ANOVA. Again, there was a significant interaction effect between participant group (HD vs controls) and cue type (gain, loss, neutral) *(Greenhouse Geisser F_1.7, 68_ = 3.46, p = 0.04, partial eta^2^ = 0.08)*. Main effects for group and cue type were not significant *(group F_1, 40_ = 0.37, p = 0.54, partial eta^2^ = 0.009; cue type F_2, 39_ = 1.60, p = 0.22, partial eta^2^ = 0.07)*.

Post-hoc analyses of the interaction revealed no significant differences between the cue types according to group (gain cue mean difference = −0.53, *p* = 0.14; loss cue mean difference = −0.33, *p* = 0.40; and neutral mean difference = 0.26, *p* = 0.51). Pairwise comparisons showed that controls’ CNV in response to gain cues was significantly greater than their response to neutral cues (gain vs neutral mean difference = −0.64, *p* = 0.03). By contrast, controls’ CNV in response to gain cues did not differ significantly from their CNV following loss cues (gain vs loss mean difference = −0.27, *p* = 0.36), and differences between controls’ CNV following loss relative to neutral cues was also non-significant (loss vs neutral mean difference = −0.37, *p* = 0.45). There were no significant differences in CNV amplitude across the three cue types within the HD group. Refer to Figure 4.

**Figure 4.**
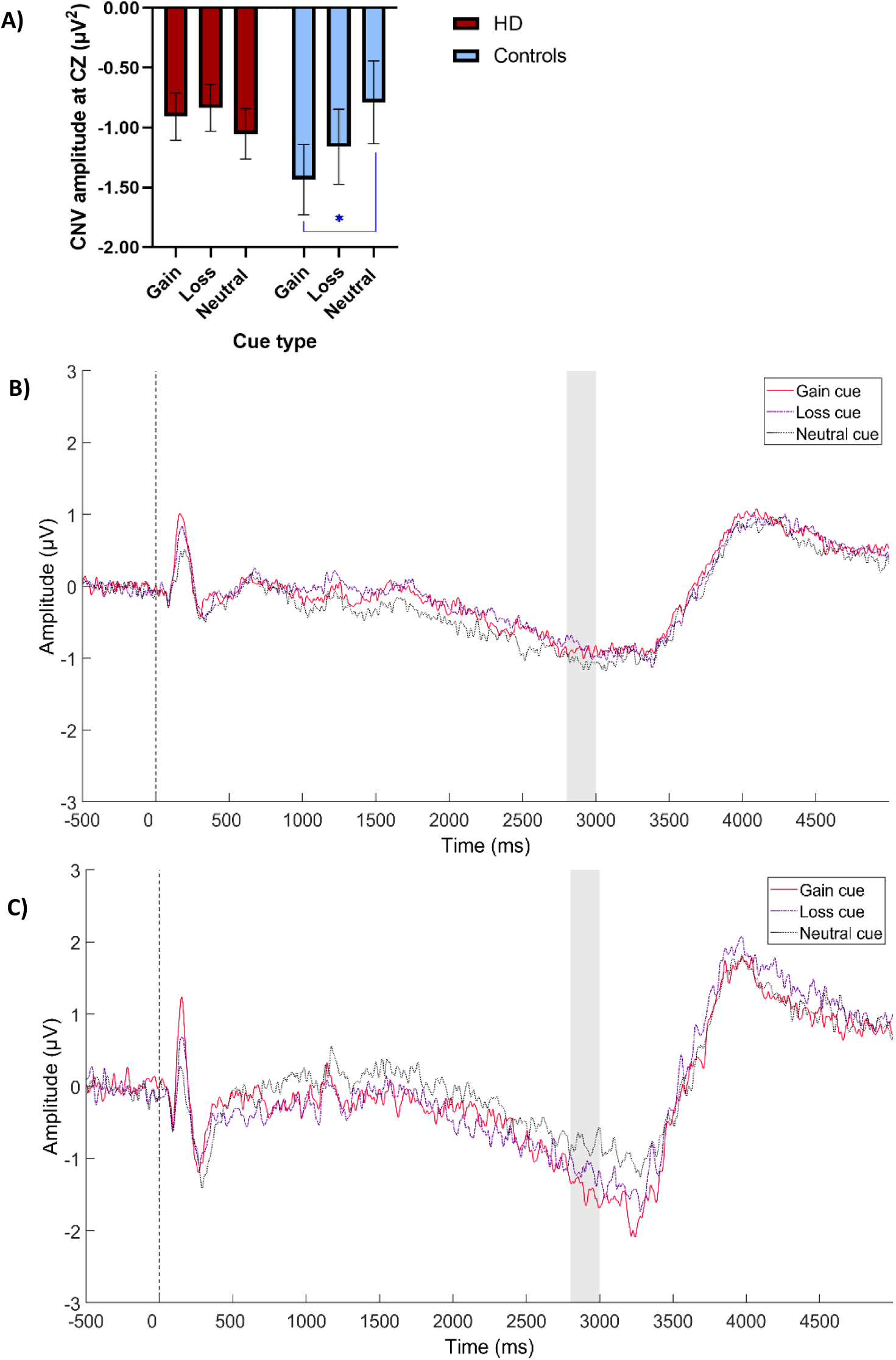
A) CNV amplitude at electrode CZ for each group averaged across trials for the different cue types (error bars reflect standard error). B) ERP plot of CNV at electrode CZ in response to each cue type for the HD group. C) ERP plot of CNV at electrode CZ in response to each cue type for the neurotypical control group. Grey bar indicates time window used for statistical analysis.

### Difference waves

To isolate amplitude elicited by the motivationally salient cues and increase statistical power for subsequent analyses, difference waves were created by subtracting the amplitude associated with neutral cues from that associated with gain and loss cues, herein referred to as Gain-P3_diff_, Loss-P3_diff_, Gain-CNV_diff,_ and Loss-CNV_diff_.

Independent t-tests and a Mann-Whitney U-test were conducted to determine group differences in these four difference waves. The Gain-P3_diff_ was of significantly greater amplitude in the control group than the HD group (Control M(SD) = −0.79(1.01); HD M(SD) = −0.17(0.39); *t_24_ = −2.58, p = 0.01*), as was the Gain-CNV_diff_ (Control M(SD) = 0.64(1.38); HD M(SD) = −0.14(0.74); *U_42_ = 138.00, p = 0.03*). The Loss-P3_diff_ and Loss-CNV_diff_, however, did not differ significantly between groups (Loss-P3_diff_ Control M(SD) = −0.36(0.69); HD M(SD) = −0.03(0.39); *t_29_ = −1.88, p = 0.07*; and Loss-CNV_diff_ Control M(SD) = 0.37(1.37); HD M(SD) = −0.22(0.83); *t_40_ = 1.69, p = 0.10*).

### Correlations between ERP difference waves, apathy, and processing speed

The difference waves that successfully discriminated between the groups (i.e., Gain-P3_diff_ and Gain-CNV_diff_) were correlated with the AES-C and SDMT. Given the exploratory goals of the correlations, no further adjustments were made for multiple comparisons (Greenland 2021). There was one extreme outlier (i.e., a Z score >±3.0) on the AES-C in the HD group, which was winsorized (i.e., 90^th^ percentile method) prior to further analysis. Non-parametric Spearman’s correlations were conducted to accommodate non-normally distributed variables where required. The results for each group are presented in Table 2.

**Table 2.**
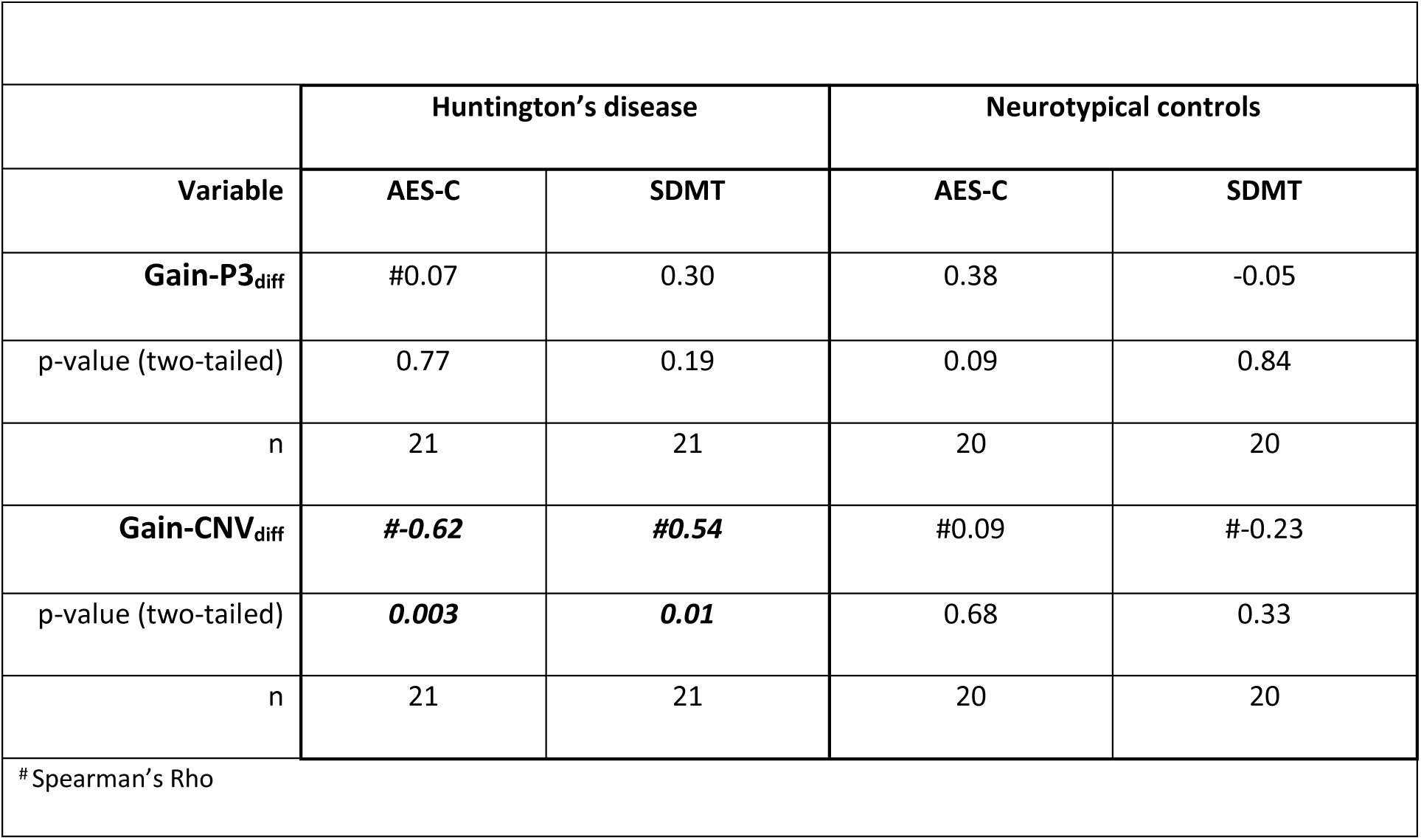
Correlations between the significant difference waves, AES-C and SDMT within each group.

|  |  |  |  |  |
| --- | --- | --- | --- | --- |
|  | Huntington's disease |  | Neurotypical controls |  |
| Variable | AES-C | SDMT | AES-C | SDMT |
| **Gain-P3<sub>diff</sub>** | #0.07 | 0.30 | 0.38 | -0.05 |
| p-value (two-tailed) | 0.77 | 0.19 | 0.09 | 0.84 |
| n | 21 | 21 | 20 | 20 |
| **Gain-CNV<sub>diff</sub>** | **#-0.62** | **#0.54** | #0.09 | #-0.23 |
| p-value (two-tailed) | **0.003** | **0.01** | 0.68 | 0.33 |
| n | 21 | 21 | 20 | 20 |
| # Spearman's Rho |  |  |  |  |

Within the HD group, there were statistically significant relationships between Gain-CNV_diff_ and scores on the AES-C and SDMT. Gain-CNV_diff_ scores were significantly *negatively* associated with AES-C scores (*rho = −0.62. n = 21, p = 0.003*), indicating that participants with less amplitude in their CNV following gain relative to neutral cues had higher apathy. By contrast, Gain-CNV_diff_ scores were significantly *positively* associated with SDMT scores (*rho = 0.54, n = 21, p = 0.01*), indicating that HD participants with greater amplitude in their CNV following gain relative to neutral cues had faster processing speeds (refer to Figure 5 for scatterplots of the significant correlations). The correlations were unchanged when conducted on the dataset that included the extreme outlier (i.e., pre-winsorization) (refer to Supplementary Material Table s1). There were no statistically significant correlations identified within the neurotypical control group.

**Figure 5.**
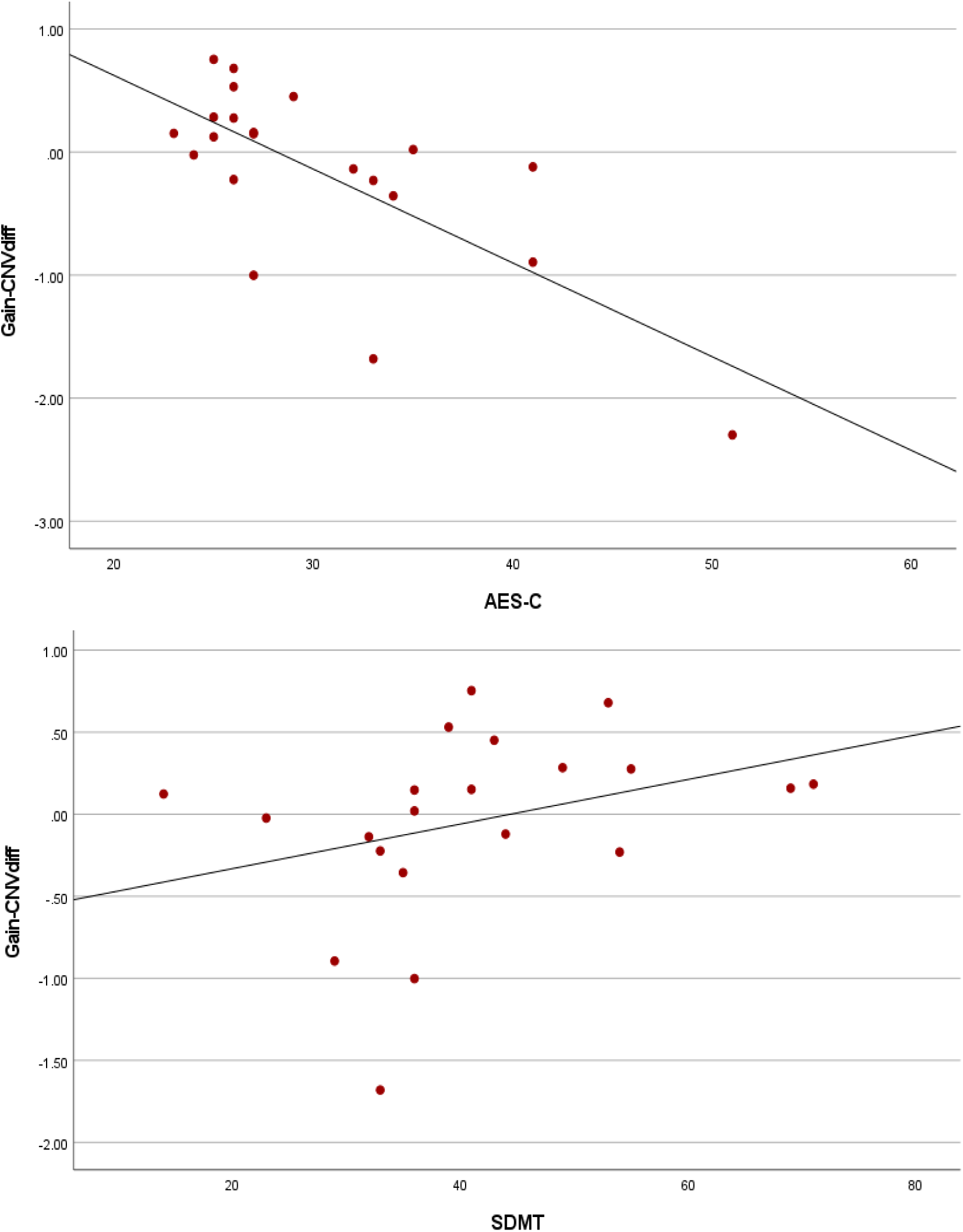
Scatterplots of correlations between Gain-CNV_diff_ and the AES-C and SDMT in the HD group.

## Discussion

In this investigation of the EEG version of the MID as a potential marker of apathy in HD, HD participants demonstrated similar P3 and CNV amplitudes in response to motivationally salient as well as neutral cues. Consistent with previous research, neurotypical controls demonstrated significantly greater P3 and CNV amplitudes in response to motivationally salient relative to neutral cues. Difference waves were calculated for gain-P3, loss-P3, gain-CNV and loss-CNV to isolate amplitude specific to the motivationally salient cues and increase statistical power. Only the gain-P3 and gain-CNV difference waves significantly differed between the groups and were included in subsequent exploratory correlations with clinical measures of apathy and processing speed.

The gain-CNV difference wave was significantly negatively associated with apathy and positively associated with processing speed in the HD group. HD participants with lower CNV amplitudes following gain relative to neutral cues had higher apathy scores, while HD participants with greater CNV amplitudes following gain relative to neutral cues had faster processing speed scores. There were no significant correlations between the difference waves and these measures within the control group.

Behaviourally, the HD group was significantly slower than the control group across all three cue types. Neither group demonstrated significantly faster response times following motivationally salient cues relative to neutral cues, which conflicts with previous studies of neurotypical controls. Previous studies, however, used samples far more restricted in age range (Novak and Foti 2015, Pfabigan et al. 2014, Zhang et al. 2017), resulting in more homogeneous response times (i.e., less response time variability enabling greater sensitivity to cue-related differences) (Dykiert et al. 2012). This reasoning is supported by post-hoc correlations that found age to be significantly positively correlated with response times for all cue types in the neurotypical control, but not HD group (refer to Supplementary Material table s2).

### P3 and CNV amplitudes

The differential effect of the motivationally salient cue types on P3 and CNV amplitude in neurotypical controls was generally consistent with three of the most methodologically similar studies (Novak and Foti 2015, Pfabigan et al. 2014, Zhang et al. 2017). Two of these studies found P3 amplitude in response to gain cues to be significantly greater than that in response to loss and neutral cues, the difference between loss and neutral amplitudes failing to reach significance (i.e., gain > loss = neutral) (Pfabigan et al. 2014, Zhang et al. 2017). The third study found that the P3 amplitude elicited by gain and loss cues was significantly greater than that elicited by neutral cues, but gain did not differ significantly from loss (i.e., gain = loss > neutral) (Novak and Foti 2015). Our findings differed in that our neurotypical control group demonstrated significant differences in P3 amplitude between all three cue types (i.e., gain > loss > neutral).

The aforementioned studies were more consistent with regards to their CNV findings; all three finding that gain and loss cues did not differ in their effect on CNV amplitudes, but both elicited significantly greater amplitudes than neutral cues (Novak and Foti 2015, Pfabigan et al. 2014, Zhang et al. 2017). By contrast, our neurotypical controls demonstrated significantly greater CNV amplitudes in response to gain cues relative to both loss and neutral cues; the latter amplitudes not differing significantly.

The primary methodological difference of our study was the large (18-to-70-year) age range of our participants, far greater than other studies (all of which recruited participants aged between 18 and 26 years) (Zhang et al. 2017, Pfabigan et al. 2014, Novak and Foti 2015). Functional MRI MID studies comparing young and older neurotypical adults are mixed with regards to the interaction effects of age and cue valence (gain vs loss) on the strength and patterns of activation (Samanez-Larkin et al. 2007, Spaniol et al. 2015). The differences in pattern of results in our study may therefore reflect increased variability associated with a broader age range of participants (Hill et al. 2018). Again, this reasoning is supported by additional post-hoc correlations, which revealed significant negative correlations between age and P3 amplitude for all three cue types in the control group, but no significant correlations with age for the HD group (refer to Supplementary Material tables s3 and s4).

Although there seem to be inconsistencies in the relative amplitude of the P3 and CNV in response to loss cues, gain cues have reliably elicited greater P3 and CNV amplitude relative to neutral cues across all these studies, including the present study. As such, gain cues may be the most reliable means of activating CBGT networks in neurotypical controls.

### Difference waves

Difference waves allowed us to quantify ERP amplitude purely in response to the two motivationally salient cues. To our knowledge, only two other studies using the EEG version of the MID have calculated difference waves, and both were for neurotypical controls (Hill et al. 2018, Novak and Foti 2015). Both studies, however, combined responses to gain and loss cues prior to subtracting the amplitude of neutral cues (Hill et al. 2018, Novak and Foti 2015). We chose to investigate the cues separately based on the study by Enzi and colleagues (2012) that found people with HD demonstrated differential patterns of activation on fMRI in response to gain and loss cues, indicating that the two motivationally salient cue types may not be interchangeable in the context of HD. Meta-analyses of the fMRI version of the MID in neurotypical controls also indicate differential activation in response to gain versus loss cues (Oldham et al. 2018, Wilson et al. 2018). This choice appears to have been justified given that only the P3 and CNV difference waves related to gain cues successfully differentiated between our HD and neurotypical control groups.

### Correlations

The significant association between gain-CNV difference wave and apathy scores in the HD group aligned with our hypothesis, and suggests that isolated CNV amplitude attributable to gain cues decreases as apathy increases in people with HD. Although not elicited using the MID, reduced CNV and P3 amplitudes have been identified in studies of people with HD in the context of other tasks (e.g., go/no-go paradigm) (Beste et al. 2008, De Tommaso et al. 2007, Turner et al. 2015).

There is also recent evidence indicating that the CNV elicited during the MID is sensitive to both motivational cue salience as well as effort discounting (Zhang and Zheng 2022). Using a modified EEG version of the MID with an additional manipulation of effort levels, Zhang and Zheng (2022) found that CNV amplitude still varied according to cue motivational salience but was reduced for both motivationally salient and neutral cues in the high effort relative to low effort condition, indicative of effort discounting. Complementary evidence for the role of effort discounting as a neurocognitive mediator of apathy in HD comes from Atkins and colleagues (2020), who found that people with premanifest HD demonstrated greater effort discounting on a cognitive task than neurotypical controls. They also found that significant effort discounting on a physical task predicted greater apathy scores on the self-report version of the AES and the Initiation subscale of the Dimensional Apathy Scale (Atkins et al. 2020, Radakovic and Abrahams 2014).

Processing speed is a highly sensitive marker of cognitive change in HD and decreases with disease progression, while apathy increases with disease progression (Paulsen et al. 2017, Thompson et al. 2012, Stout, Andrews, and Glikmann-Johnston 2017, Stout et al. 2014). Therefore, the significant positive correlation between gain-CNV difference wave and processing speed scores complements the negative correlation between gain-CNV difference wave and apathy scores.

The absence in our data of significant correlations between gain-P3 difference wave, apathy, and processing speed scores in either group was unexpected. This may reflect the broader functional significance of the P3 relative to the CNV (Glazer et al. 2018, Luck 2014, Polich 2012). The absence of significant correlations between the difference waves and apathy and processing speed scores in the control group may also be explained by the reduced spread of scores and therefore limited sensitivity of these clinical measures in neurotypical participants.

Of primary interest to us were the *reduced* ERP amplitudes and inferred *lack* of CBGT activation in response to motivationally salient cues in participants with HD. Motivationally salient cue processing is considered one of the neurocognitive subprocesses underpinning motivated, goal-directed behaviour (Chong 2018, Le Heron et al. 2018, Morris, O’Callaghan, and Le Heron 2022). In the EEG version of the MID, motivationally salient cue processing is measured by the P3, which is functionally related to the allocation of attentional resources in the service of memory, but also modulated by the dopaminergic and noradrenergic systems (Huang, Chen, and Zhang 2015, Polich 2012). The lack of P3 sensitivity to motivational salience in HD participants may reflect a deficit in immediate encoding of reward value by dopaminergic neurons (Bromberg-Martin, Matsumoto, and Hikosaka 2010, Saunders et al. 2018). Previous research also found that people with HD did not demonstrate the expected differential skin conductance responses to wins and losses on a gambling task; further evidence of insensitivity to motivational salience in people with HD (Campbell, Stout, and Finn 2004). Failure to encode the relative motivational salience of the different cue types would, in turn, fail to modulate preparatory responses across cue types and result in uniformity of CNV amplitude, as appeared to be the case for the HD group in the present study.

At a broader neuroanatomical level, these findings align with the growing consensus that apathy in HD reflects CBGT network disruption, likely beginning in the striatum (Atkins et al. 2020, Enzi et al. 2012, Lanctôt et al. 2017, Le Heron et al. 2018, Morris, O’Callaghan, and Le Heron 2022). How striatal and broader CBGT structural changes interact with dopamine availability to affect apathy in HD requires further investigation. There is a high prevalence of apathy in brain disorders with deficient as well excess cortico-striatal dopamine availability (Epstein and Silbersweig 2015), consistent with a proposed “inverted U-shape function, where too much or too little dopamine impairs performance” (Cools and D’Esposito 2011, 114).

Dopamine availability also changes with HD progression and naturally interacts with the dynamics of other neurotransmitter systems (André, Cepeda, and Levine 2010, Schwab et al. 2015). Given the sensitivity of ERPs to acute pharmacological manipulation of neurotransmitters (Linssen et al. 2011, Schutte et al. 2020), the EEG version of the MID would be well suited to further investigation of the role of dopamine in HD-related apathy.

From a treatment development perspective, understanding and treating apathy and other non-motor symptoms in HD requires measures that are conceptually and clinically relevant, and able to elicit measurable physiological changes in neuroanatomical regions of interest. The current study provides preliminary evidence supporting the EEG version of the MID as a proxy measure of apathy in HD. In addition to cost-effectively assaying neuropsychological phenomena, markers such as those generated by the EEG version of the MID, are sensitive to a variety of treatment types (e.g., medication, brain stimulation, psychological and behavioural therapies) with fewer confounds than those associated with cognitive and behavioural measures (Ahn et al. 2019, Alhaj, Wisniewski, and McAllister-Williams 2011, Briels et al. 2020). This is a particularly important consideration for proof-of-concept and phase I clinical trials where functional or clinical changes are not expected but some evidence of biological target engagement may still be required (e.g., Hua et al. 2022).

### Limitations

These findings require replication in a larger sample of people with HD using more complex statistical analyses that can control for covariates such as age, education, and phenomena related to apathy (e.g., depression and fatigue). Ideally, the potential confounding effect of dopamine antagonists (i.e., risperidone and tetrabenazine) on the amplitude of ERPs should also be controlled in future investigations. Nevertheless, we do note that Enzi and colleagues (2012) found that exclusion of HD participants on medication did not alter their results using the fMRI version of the MID, and that P3 and CNV amplitude have been reduced in previous studies of people with HD who were medication free (Beste et al. 2008, De Tommaso et al. 2007), therefore we do not suspect that medication use would have accounted for the current findings.

## Conclusion

Non-motor symptoms of HD are associated with significant functional burden and reduced quality of life (Fritz et al. 2018, Jacobs, Hart, and Roos 2018, Martinez-Horta et al. 2016, Paoli et al. 2017). Neuropsychological symptoms such as apathy are also multidetermined and difficult to measure with the sensitivity and specificity required for the development of novel treatments. These findings suggest that the EEG version of the MID may be used as a conceptually valid, proxy measure of apathy in HD.

## Data Availability

All data produced in the present study are available upon reasonable request to the authors from genuine researchers who agree to maintain the confidentiality of the data.

## Acknowledgements

MCD was supported by the Research Training Program Stipend and a Monash Graduate Excellence Scholarship. KEH was supported by National Health and Medical Research Council (NHMRC) Fellowships (1082894 and 1135558). PBF was supported by an Investigator Fellowship from the NHMRC (1193596). ATH was supported by an Alfred Deakin Postdoctoral Research Fellowship. We gratefully acknowledge the time, involvement, and feedback from the participants. This research would not have been possible without them. We also acknowledge the assistance with participant recruitment provided by the Statewide Progressive Neurological Disease Service at Calvary Health Care Bethlehem.

## Supplementary Material

**Table s1.** Correlations between the significant difference waves and AES-C with the extreme outlier (i.e., prior to winsorization) in the HD group.

| Variable | Gain-P3<sub>diff</sub> | Gain-CNV<sub>diff</sub> |
| --- | --- | --- |
| **AES-C** | #0.07 | **#-0.62** |
| p-value (two-tailed) | 0.78 | **0.003** |
| n | 21 | 21 |
| #Spearman's Rho |  |  |

**Table s2.** Correlations between the response times (RT) for each cue type and age within each group.

| Variable | Huntington's disease |  |  | Neurotypical controls |  |  |
| --- | --- | --- | --- | --- | --- | --- |
|  | Gain RT | Loss RT | Neutral RT | Gain RT | Loss RT | Neutral RT |
| **Age** | 0.30 | 0.31 | 0.32 | **0.53** | **#0.50** | **0.48** |
| p-value (two-tailed) | 0.20 | 0.16 | 0.15 | **0.02** | **0.03** | **0.03** |
| n | 22 | 22 | 22 | 20 | 20 | 20 |
| #Spearman's Rho |  |  |  |  |  |  |

**Table s3.** Correlations between the P3 and CNV amplitudes for each cue type and age within the control group.

| Variable | Gain P3 | Loss P3 | Neutral P3 | Gain CNV | Loss CNV | Neutral CNV |
| --- | --- | --- | --- | --- | --- | --- |
| **Age** | **-0.68** | **-0.57** | **-0.58** | -0.44 | -0.27 | #0.06 |
| p-value (two-tailed) | **<0.001** | **0.009** | **0.007** | 0.051 | 0.25 | 0.79 |
| n | 20 | 20 | 20 | 20 | 20 | 20 |
| #Spearman's Rho |  |  |  |  |  |  |

**Table S4.** Correlations between the P3 and CNV amplitudes for each cue type and age within the HD group.

|  |  |  |  |  |  |  |
| --- | --- | --- | --- | --- | --- | --- |
| **Variable** | **Gain P3** | **Loss P3** | **Neutral P3** | **Gain CNV** | **Loss CNV** | **Neutral CNV** |
| **Age** | #0.06 | 0.10 | -0.10 | -0.17 | 0.13 | -0.19 |
| p-value (two-tailed) | 0.78 | 0.66 | 0.64 | 0.45 | 0.58 | 0.40 |
| n | 22 | 22 | 22 | 22 | 22 | 22 |
| #Spearman's Rho |  |  |  |  |  |  |

